# *De novo FZR1* loss-of-function variants cause developmental and epileptic encephalopathies including Myoclonic Atonic Epilepsy

**DOI:** 10.1101/2021.06.12.21256778

**Authors:** Sathiya N. Manivannan, Jolien Roovers, Noor Smal, Candace T. Myers, Dilsad Turkdogan, Filip Roelens, Oguz Kanca, Hyung-Lok Chung, Tasja Scholz, Katharina Hermann, Tatjana Bierhals, S. Hande Caglayan, Hannah Stamberger, Heather Mefford, Peter de Jonghe, Shinya Yamamoto, Sarah Weckhuysen, Hugo J. Bellen

## Abstract

*FZR1*, which encodes the Cdh1 subunit of the Anaphase Promoting Complex, plays an important role in neurodevelopment by regulating cell cycle and by its multiple post-mitotic functions in neurons. In this study, evaluation of 250 unrelated patients with developmental epileptic encephalopathies (DEE) and a connection on GeneMatcher led to the identification of three *de novo* missense variants in *FZR1*. Two variants led to the same amino acid change. All individuals had a DEE with childhood onset generalized epilepsy, intellectual disability, mild ataxia and normal head circumference. Two individuals were diagnosed with the DEE subtype Myoclonic Atonic Epilepsy (MAE). We provide gene burden testing using two independent statistical tests to support *FZR1* association with DEE. Further, we provide functional evidence that the missense variants are loss-of*-*function (LOF) alleles using *Drosophila* neurodevelopment assays. Using three fly mutant alleles of the *Drosophila* homolog *fzr* and overexpression studies, we show that patient variants do not support proper neurodevelopment. With the recent report of a patient with neonatal-onset DEE with microcephaly who also carries a *de novo FZR1* missense variant, our study consolidates the relationship between *FZR1* and DEE, and expands the associated phenotype. We conclude that heterozygous LOF of *FZR1* leads to DEE associated with a spectrum of neonatal to childhood onset seizure types, developmental delay and mild ataxia. Microcephaly can be present but is not an essential feature of *FZR1*-encephalopathy. In summary, our approach of targeted sequencing using novel gene candidates and functional testing in *Drosophila* will help solve undiagnosed MAE/DEE cases.

## Introduction

Developmental and Epileptic Encephalopathies (DEEs) are a heterogeneous group of disabling disorders, characterized by a combination of severe epilepsy and neurodevelopmental problems^1^. Seizures in DEE patients usually have an early age-of-onset and are often refractory to anti-epileptic drugs. The majority of DEE cases are thought to have a genetic basis, usually in the form of rare *de novo* variants with a highly disruptive effect on gene expression or protein function. Pathogenic variants in more than a hundred genes have been associated with a DEE phenotype so far^2-4^. Identifying the causal variant in a DEE patient can bring an end to an often long and stressful diagnostic odyssey and may lead to improved or targeted treatment for a subset of the cases^2; 5; 6^.

We set out to identify novel variants in DEE patients by screening a large cohort of patients with Myoclonic Atonic Epilepsy (MAE), a sub-type of DEE, as part of the efforts of the EuroEPINOMICS-RES consortium^7^. MAE is characterized by generalized seizure types including myoclonic, atonic, myoclonic atonic, absence and tonic-clonic seizures. The seizure onset is typically between 7 months and 6 years^8^. Using a combination approach of trio whole-exome sequencing (WES) and subsequent screening with a targeted gene panel including candidate genes for MAE, we identified two individuals diagnosed with MAE who each carried a unique *de novo* missense variant in *FZR1*. A third individual with a DEE with childhood onset generalized epilepsy and a *de novo* missense variant was identified through GeneMatcher.

*FZR1* (*fizzy and cell division cycle 20 related 1*, MIM:603619) encodes Cdh1, one of the two regulatory subunits of the Anaphase Promoting Complex (APC) that confers the substrate-specificity on this E3 ubiquitin ligase complex^9^. APC was initially identified to play a key role in mitotic cell cycle progression^10^. When Cdh1 associates with APC (Cdh1-APC), it controls mitotic exit leading to G1 arrest^11^. *Cdh1*/*Fzr1* knockout in mice leads to embryonic lethality, suggesting that this gene plays critical roles in development *in vivo* in mammals^12^. These *Fzr1* knockout mice, which die as embryos, have impaired cortical neurogenesis due to delay of mitotic exit, leading to reduced cortical size and thickness^13^. Cdh1-APC also has a prominent function in post-mitotic cells in the nervous system^14^. It controls neuronal survival^15; 16^, axonal growth^17^, and synapse formation and function^18^. Similar to its mammalian counterpart, the *Drosophila* homolog of *FZR1, fzr* (*fizzy-related)*, has been studied in the context neurodevelopment in addition to its role in cell-cycle regulation as part of the APC^17; 19-21^. *fzr* is necessary for photoreceptor patterning and regulates glial cell migration from the brain into the eye imaginal disc^17^. Through a forward genetic screen, we previously isolated two *fzr* alleles which displayed abnormal retina pattern and defects in electroretinograms (ERG)^22^. These phenotypes have been reported in hypomorphic mutants of *fzr* which were first identified through their rough eyes, leading to one of the gene’s synonyms ‘*retina aberrant in pattern* (*rap*)’^23^. These previously generated *fzr* alleles provide genetic tools to test the functionality of the human *FZR1* variants identified in DEE patients.

During the course of our study, a different rare variant in *FZR1* was reported in an individual with microcephaly and DEE^24^. This study showed that the missense variant found in their patient led to decreased stability of the FZR1 protein in patient leukocytes and in HEK293T cells as well as accumulation of proteins targeted by Cdh1-APC^24^. The mutant protein was also unable to rescue the aberrant cell cycle distribution of primary cortical progenitors from *Cdh1/Fzr1* knockout mice. Based on these data, the authors argued that this variant potentially contributes to microcephaly in the patient.

Here, we present three individuals with DEE and a novel *de novo* missense variant in *FZR1*. These cases consolidate the role of *FZR1* in DEE and expand the phenotypic spectrum of *FZR1*-related encephalopathy to DEE with childhood onset generalized epilepsy, mild ataxia and normal head circumference. To determine the functional consequence of the missense variants, we assessed the ability of the variant proteins to rescue the eye phenotype of the fly *fzr* mutant alleles isolated from a forward genetic screen using mosaic analysis. Furthermore, we also generated a new loss-of-function allele of *fzr* using the CRIMIC technology^25^ to study the impact of the patients’ variants during development. Through these assays in *Drosophila*, we provide functional evidence for a LOF mechanism of the pathogenic variants found in our patients, further supporting the involvement of FZR1 in MAE.

## Subjects and methods

### Identification of patients with *de novo FZR1* variants

Patient 1 was recruited to the project on MAE of the EuroEPINOMICS-RES consortium^7^. WES was performed on an initial cohort of 39 parent-offspring trios (including Patient 1) with MAE at the Wellcome Trust Sanger Institute (Hinxton, Cambridgeshire). WES and subsequent data analyses were performed as previously described^26^. The GenomeComb program was used for annotation and filtering of the data^27^. Coding variants that lead to a missense change, stop gain or stop loss, frameshift or essential splicing change were retained for further filtering. Variants that were present in the ExAC^28^ and Genome Aggregation Database (gnomad, v2.1.1)^29^ with a frequency > 0.01 or > 0 for homozygous and X-chromosome or heterozygous variants respectively were excluded. We also excluded variants in genes that are not expressed in the brain in the Genotype-Tissue Expression project database (GTEx V8)^30^. Pathogenicity of missense variants was predicted using the Combined Annotation Dependent Depletion scores (CADD, v1.6)^31^ and missense variants with a CADD-score < 20 were excluded. All remaining candidate variants following a recessive, *de novo* or X-linked inheritance were validated using Sanger sequencing.

Patient 2 was part of a follow-up cohort consisting of 211 DEE patients, including 89 probands with MAE, and 122 probands with Dravet or Dravet-like syndrome. The cohort was screened using a targeted gene panel using Molecular Inversion Probes and consisted of the coding regions of 12 known and 40 candidate genes (at time of screening) for MAE, including *FZR1* (see supplemental Table 1). Candidate genes included in this panel were selected from a list of genes with single *de novo* hits identified in the EuroEPINOMICS-RES WES data. Data analysis and variant filtering was performed as previously described^32^. Segregation analysis was performed using Sanger sequencing for all nonsynonymous, frameshift, and splice-site variants that were not present in the ExAC set of ∼61,000 WES^28^. Paternity and maternity were confirmed using the Powerplex®16S system (Promega).

Patient 3 was identified through GeneMatcher^33^. Trio-WES was performed in a clinical diagnostic setting using DNA from the patient and both healthy parents. Trio-exome data were filtered for non-synonymous *de novo* variants absent in the general population (gnomAD), and for rare biallelic variants with minor allele frequency (MAF) <0.1% and absence of homozygous carriers in the aforementioned database. The functional impact of the identified missense variants was predicted using the CADD, v1.6^31^ SIFT^34^ and Polyphen^35^ pathogenicity prediction tools.

The Ethical Committee of the University of Antwerp, Belgium gave ethical approval for the study. Parents of each patient signed an informed consent form for participation in the study.

### Gene burden analysis

A gene burden analysis was performed using denovolyzeR^36^ for the two *de novo* missense variants identified in the combined research cohort of 250 patients examined in our study. In parallel, we determined the number of rare, potential damaging variants in *FZR1* in the control population set of gnomAD^28^ based on CADD^31^, SIFT^34^ and Polyphen^35^ scores using the Variant Effect Predictor tool from Ensembl^37^. We filtered missense variants and loss-of-function variants that had a general population frequency of less than or equal to 0.01, had a CADD score greater than or equal to 20, and were predicted to be Pathogenic/Likely pathogenic by SIFT and Polyphen. We performed a chi-square test to evaluate the allele-count burden as well as number of unique variant burden in the DEE patient cohorts compared to the gnomAD (control) population.

### Evaluation of relative positions of affected residues in the 3D structural model of human Cdh1

To examine the localization of the residues predicted to be altered by the patients’ missense variants, we used the 3D structural model of Cdh1 generated from an electron microscopy reconstruction of the *Homo sapiens* APC (PDB ID:4ui9)^9^. We used PyMOL (Molecular Graphics System, Version 2.0 Schrödinger, LLC). to map the location of the residues affected. We also predicted the location of the *Drosophila* Fzr^A^ variant based on the homology of human FZR1 and *Drosophila* Fzr proteins (Figure S2A).

### Functional assessment of DEE-associated missense variants in *Drosophila melanogaster*

The molecular lesions of two EMS-induced variants identified from our previous forward genetic screen^*22*^, *fzr*^*A*^ and *fzr*^*B*^, were identified using Sanger sequencing of PCR amplicons of genomic DNA^38^. ERG analysis on mosaic adult eye were performed as described earlier^39^. We used CRISPR/Cas9 technology to insert a T2A-GAL4 gene trap cassette based on CRISPR mediated Integration Cassette (CRIMIC) technology into the intron of *fzr* at ChrX:4,676,058 (*Drosophila melanogaster* reference genome, Assembly 6) and generated the *fzr*^*CR00643-TG4*.*2*^ allele (referred in the text as *fzr*^*T2A-GAL4*^). Expression of GAL4 from the *fzr*^*T2A-GAL4*^ allele was examined using a UAS-nls::RFP reporter [Bloomington Drosophila Stock Center (BDSC) stock number: 8546). Complementation tests were performed using the *fzr*^*ie28*^ allele^19^ (a kind gift from Dr. Christian Klambt) and *Dp(1;3)DC472* P[acman] clone inserted in the third chromosome at the VK33 docking site^40-42^. To perform rescue experiments of *fzr* mutants, we subcloned the full length *fzr* cDNA from a pUASt-fzr vector^19^. *Drosophila fzr* cDNA carrying the variants at positions corresponding to the human variants were generated by site-directed mutagenesis using QuickChange II kit (Agilent). As patient 2 and patient 3 carry a variant affecting the same nucleotide and leading to the same protein change, only the C>G variant of patient 2 was modeled. Wild-type (wt) and mutant *fzr* expressing UAS constructs were integrated into VK33 docking site on the fly third chromosome via phiC31 transgenics^40^. Transgenic lines were recovered and established based on standard crossing schemes^43^. Rescue experiments for eye phenotypes of *fzr*^*B*^ mutants were performed using the MARCM (Mosaic analysis with a repressible cell marker) technique^44^. MARCM-based rescue was achieved by crossing flies of the following genotype: *y w fzr*^*B*^, *FRT19A/FM7c, Kr-GAL4, UAS-GFP* (BDSC 52385) and *tub-GAL80, ey-FLP, FRT19A/*Y; *Act-GAL4, UAS-GFP/CyO* (FRT19A tester line). The FRT19A tester line was generated by crossing the following stocks *tub-Gal80, ey-FLP, FRT19A* (BDSC 42717) and *FRT19A; Act5C-GAL4, UAS-GFP/CyO* (BDSC 42726). Rescue of the lethality of the *fzr*^*T2A-GAL4*^ allele was assessed based on the T2A-GAL4 strategy^25; 45^, and achieved by setting the following cross: *fzr*^*T2A-GAL4*^ /FM7, *Act-GFP* crossed to *UAS-fzr* (*wt* or *variant*) and evaluation of non-GFP male larvae at late third instar stage. Overexpression studies in the developing eye imaginal disc were carried out using *ey3*.*5-GAL4* (*y*^*1*^ *w*^*1118*^ ; *P{ey3*.*5-GAL4*.*Exel}2*) (BDSC: 8220)^17^.

### Electroretinogram analysis of adult mosaic eyes

*y w fzr*^*B*^, *FRT19A/FM7c, Kr-GAL4, UAS-GFP and y w fzr*^*A*^, *FRT19A/FM7c, Kr-GAL4, UAS-GFP* (BDSC 52384) females were crossed with *y w FRT19A; ey-FLP* (BDSC 5579). The resulting females carried mosaic eyes with mutants identifiable with the lack of red pigmentation. ERG analysis on mosaics were performed as described earlier^22; 39^.

### Immunofluorescence analysis of *Drosophila* tissue

Immunofluorescence analysis was performed as described earlier on embryos and third instar larval eye imaginal discs^17; 19^. The following antibodies from Developmental Studies Hybridoma Bank (University of Iowa) or commercial sources were used: mouse anti-Repo (1:50, 8D12)^46^, rat anti-ELAV (1:50, 7E8A10)^47^, rabbit anti-HRP (1:50, Jackson ImmunoResearch, #323-005-021), chicken anti-GFP (1:1000, Abcam ab13970), rabbit anti-mCherry (1:500, Abcam ab167453), Alexa488 conjugated goat anti-Chicken (1:200, ThermoFisher A-11039), Alexa647 conjugated goat anti-mouse (1:250, Abcam ab150115), Alexa647 conjugated goat anti-rat (1:250, Abcam ab150159), Alexa594 conjugated goat anti-rabbit (1:1000, ThermoFisher A-11012), Alexa405 conjugated goat anti-rabbit (1:100, ThermoFisher A-31556). Samples were imaged using a LSM 500 confocal microscope (Zeiss) to generate a Z-stack of different focal planes. The confocal Z-stack images were used to generate maximum intensity Z-projection using ImageJ^48^.

### Mammalian overexpression vectors, cell culture, Immunofluorescence, and western blot

Human *FZR1* cDNA (NM_001136198.1) in the donor vector pDONR221 (HsCD00042756;) was purchased from Harvard cDNA clone repository. The coding region was cloned into the pEzy-eGFP (Addgene #18671) destination vector in frame with an N-terminal eGFP tag using Gateway™ LR Clonase™ II Enzyme mix (ThermoFisher 11791020) following the manufacturer’s recommended protocol. The variants observed in the DEE patients were introduced into the pEzy-eGFP-FZR1 using Quickchange II site directed mutagenesis kit (Agilent).

Human embryonic kidney cells (HEK293) were grown in Dulbecco’s Modified Eagle’s Medium (DMEM; Thermofisher) supplemented with 10% fetal bovine serum. Cells were transfected with overexpression vectors using Lipofectamine 3000 reagent (ThermoFisher) with OptiMEM media (Thermofisher) using the manufacturer’s recommended protocol. Cells were fixed using 4% Paraformaldehyde in PBS at room temperature. Immunofluorescence staining was performed using chicken anti-GFP antibody (1:1000, Abcam ab13970) as described earlier^49^. Nucleus and actin-cytoskeleton were visualized using DAPI (4’,6-diamidino-2-phenylindole, ThermoFisher D1306) and Alexa647 conjugated Phalloidin (Cell signaling, #8940) respectively. Cells were imaged using Zeiss LSM 500 confocal microscope. A 2-micron optical section with the nucleus in focus were obtained. Protein isolation and western blot analysis was performed 36 hours post transfection as described earlier^49^. Chicken anti-GFP (1:1000, Abcam ab13970) and IRDye® 800CW donkey anti-chicken Secondary Antibody (LiCOR P/N: 926-32218) were used for detecting N-terminal GFP tagged FZR1 protein.

### Primers used in the study

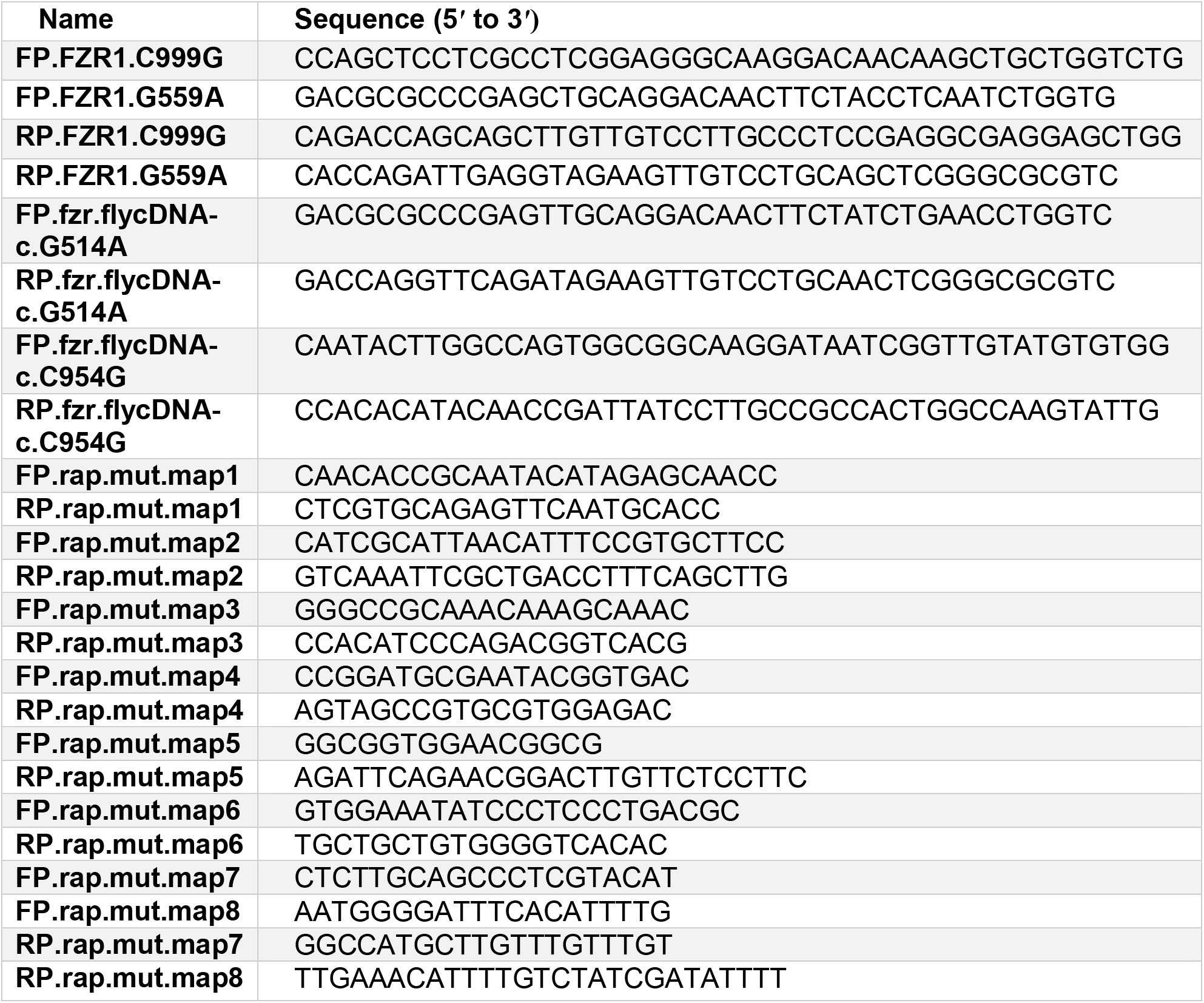

## Results

### Clinical case reports

A summary of the clinical history of the three patients described above, and the patient described by Rodriguez et al.^24^ can be found in Table 1. In compliance with the privacy rules of MedRxiv, the details of patient’s symptoms, genetic diagnosis and treatment information are available to readers upon a reasonable request to the corresponding author.

**Table 1.**
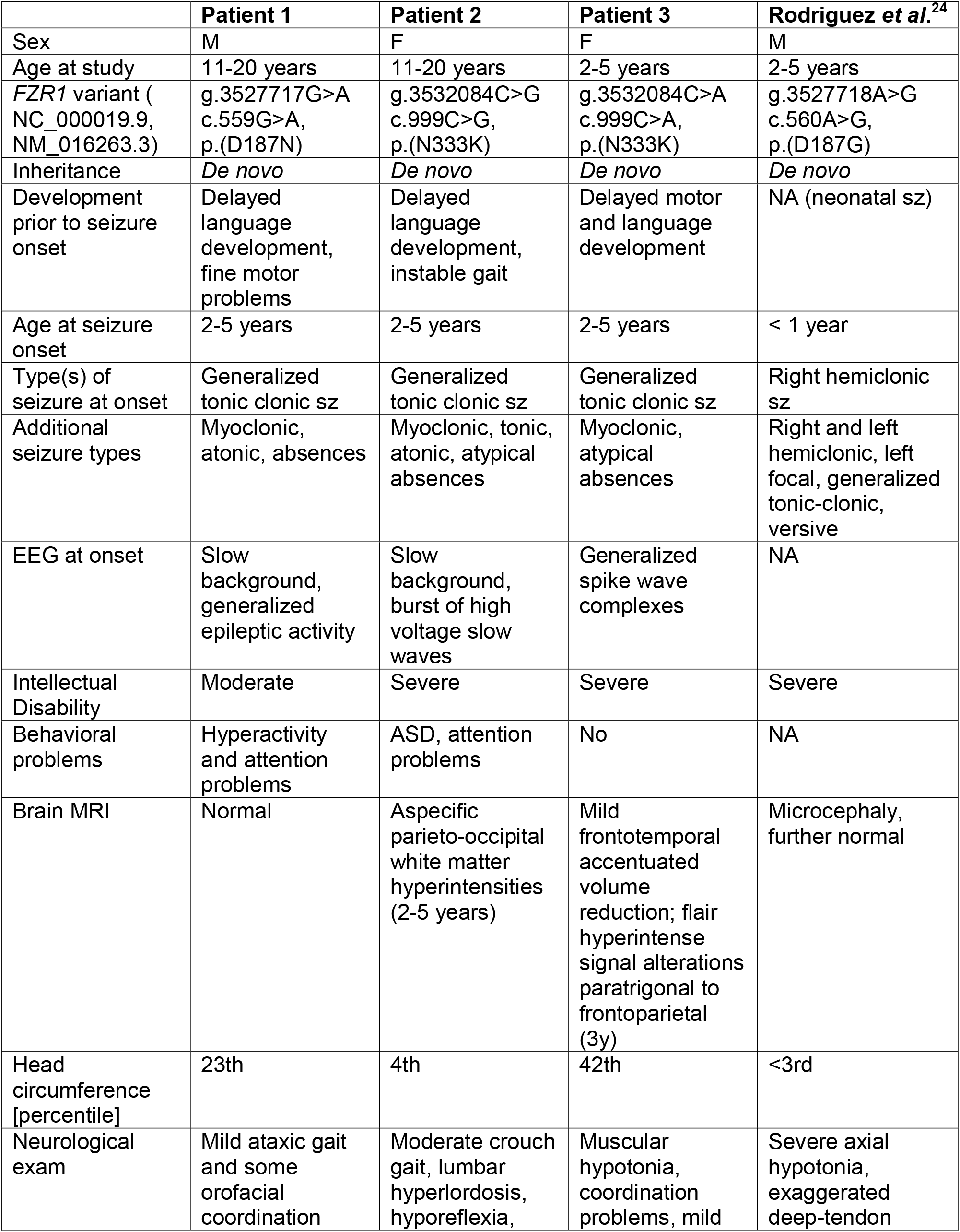
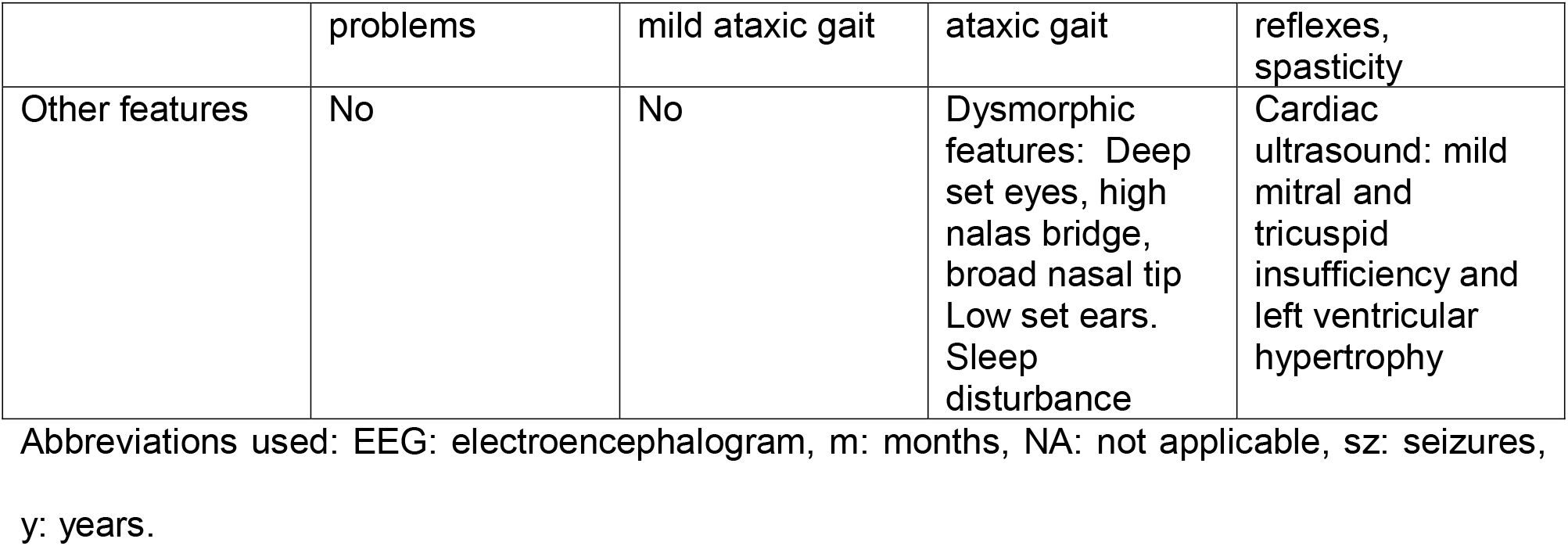
Clinical features.

|  | **Patient 1** | **Patient 2** | **Patient 3** | **Rodriguez et al.<sup>24</sup>** |
| --- | --- | --- | --- | --- |
| Sex | M | F | F | M |
| Age at study | 11-20 years | 11-20 years | 2-5 years | 2-5 years |
| *FZR1* variant (NC_000019.9, NM_016263.3) | g.3527717G>A<br>c.559G>A,<br>p.(D187N) | g.3532084C>G<br>c.999C>G,<br>p.(N333K) | g.3532084C>A<br>c.999C>A,<br>p.(N333K) | g.3527718A>G<br>c.560A>G,<br>p.(D187G) |
| Inheritance | *De novo* | *De novo* | *De novo* | *De novo* |
| Development prior to seizure onset | Delayed language development, fine motor problems | Delayed language development, instable gait | Delayed motor and language development | NA (neonatal sz) |
| Age at seizure onset | 2-5 years | 2-5 years | 2-5 years | < 1 year |
| Type(s) of seizure at onset | Generalized tonic clonic sz | Generalized tonic clonic sz | Generalized tonic clonic sz | Right hemiclonic sz |
| Additional seizure types | Myoclonic, atonic, absences | Myoclonic, tonic, atonic, atypical absences | Myoclonic, atypical absences | Right and left hemiclonic, left focal, generalized tonic-clonic, versive |
| EEG at onset | Slow background, generalized epileptic activity | Slow background, burst of high voltage slow waves | Generalized spike wave complexes | NA |
| Intellectual Disability | Moderate | Severe | Severe | Severe |
| Behavioral problems | Hyperactivity and attention problems | ASD, attention problems | No | NA |
| Brain MRI | Normal | Aspecific parieto-occipital white matter hyperintensities (2-5 years) | Mild frontotemporal accentuated volume reduction; flair hyperintense signal alterations paratrigonal to frontoparietal (3y) | Microcephaly, further normal |
| Head circumference [percentile] | 23th | 4th | 42th | <3rd |
| Neurological exam | Mild ataxic gait and some orofacial coordination | Moderate crouch gait, lumbar hyperlordosis, hyporeflexia, | Muscular hypotonia, coordination problems, mild | Severe axial hypotonia, exaggerated deep-tendon |

|  | problems | mild ataxic gait | ataxic gait | reflexes, spasticity |
| --- | --- | --- | --- | --- |
| Other features | No | No | Dysmorphic features: Deep set eyes, high nasal bridge, broad nasal tip<br>Low set ears.<br>Sleep disturbance | Cardiac ultrasound: mild mitral and tricuspid insufficiency and left ventricular hypertrophy |

Abbreviations used: EEG: electroencephalogram, m: months, NA: not applicable, sz: seizures, y: years.

WES on patient 1 and unaffected parents yielded a *de novo* missense variant in *FZR1* [c.559G>A, p.(D187N), Table 1] located at the same residue as the previously reported *de novo* missense variant in a patient with microcephaly, psychomotor retardation, and epilepsy [p.(D187G)]^24^. No other variants passed the filtering steps as described in the methods section. A relative with mild learning disabilities but no seizures did not carry the variant. The variant is not present in the gnomAD database and has a CADD score of 29.7^31^, SIFT-score of 0.01,^34^ and PolyPhen score of 0.936^35^, all indicating a deleterious or probably damaging effect. According to the gnomAD database^29^, *FZR1* has a pLI score of 1, o/e score of 0.04, and a z-score of 3.64 for missense variants, showing that this gene is intolerant to both LOF (nonsense, frameshift, core splicing) and missense variants.

Following the identification of a *de novo FZR1* missense variant in Patient 1, a follow-up panel-screening resulted in the identification of another *de novo FZR1* missense variant [c.999C>G, p.(N333K), Table 1] in Patient 2. This variant had a CADD score of 23.6, SIFT-score of 0.01 and PolyPhen score of 0.814, all predicting a deleterious or possibly damaging effect. This variant was also absent from the gnomAD database. Previous genetic tests on this patient included karyotype analysis and *SCN1A* and *PCDH19* screening, all of which were negative. Array-based comparative genomic hybridization (array-CGH) showed two duplications on Chr11p11.2. These copy number variations were inherited from the healthy mother and did not include disease-associated genes. Therefore, they deemed non-contributing to the patient phenotype.

Patient 3 was identified through Genematcher, and carries a *de novo* missense variant at the same nucleotide position as patient 2, leading to an identical amino acid change [c.999C>A, p.(N333K), table 1], with identical CADD, SIFT and PolyPhen scores. Previous chromosome analysis and array-CGH were normal. Filtering of exome data retained homozygous variants in two additional genes, not yet reported in the context of rare Mendelian disorders. The missense variant c.2105_2106delinsTC, p.(S702F) in *PTPN21* (NM_007039.4) had a SIFT-score of 0.000 and PolyPhen score of 0.996, predicting a probably damaging effect. This multinucleotide variant is present twice heterozygous in gnomAD. PTPN21 (Tyrosine-protein phosphatase non-receptor type 21, MIM: 603271) is an oncogenic protein known to be upregulated in several types of cancer cells^50^ and functions as a key regulator of inflammation^51^. Two non-synonymous single-nucleotide polymorphisms in PTPN21 showed association to schizophrenia in a GWAS study^52^. In neurons, PTPN21 controls the activity of KIF1C, a fast organelle transporter implicated in the transport of dense core vesicles and the delivery of integrins to cell adhesions^53^. It was further shown to positively influence cortical neuronal survival and to enhance neuritic length^54^. The second homozygous variant identified in patient 3 was the truncating variant c.577C>T, p.(R193*) in the last exon of *TPD52L2* (NM_199360.3). TPD52L2 (Tumor protein D52-like 2, MIM: 603747) is an ubiquitously expressed tumor protein shown to be involved in multiple membrane trafficking pathways, and to affect cell proliferation, adhesion and invasion^55,56^. As the *de novo FZR1* missense variant resulted in the same amino-acid substitution as identified in patient 2 who has a very similar phenotype, we concluded that the *FZR1* variant is most likely to underlie the neurological disease of patient 3. We can however not totally exclude the possibility that the variants in *PTPN21* and *TPD52L2* contribute to the phenotype of the patient 3.

Having found these cases, we tested the association between rare variants in *FZR1* and DEE using two different gene burden analysis tools. First, we used the tool denovolyzeR^36^ which evaluates the overrepresentation of *de novo* variants that affect the protein sequence (missense and loss of function variants) in a specific gene. We found that identification of two *de novo* variants in *FZR1* amongst the 250 patients in our research cohort is a 212-fold enrichment (Poisson distribution P-value of 4.41e-05) over the expected number of *de novo* variants, suggesting a strong association of *FZR1 de novo* missense variants with the disorder. Second, we compared the rate of rare, damaging variants in our research cohort (250 patients) with that of general population in gnomAD (141,456 samples)^29^ in *FZR1*. For this, we re-analyzed both the WES dataset and the targeted panel sequencing cohort of our patient population for the presence of damaging, rare variants regardless of inheritance, and did not identify any additional qualifying variants. Once again, we found a significant enrichment in the number of damaging variants in *FZR1* within our patient cohort (2/250 patients) compared to that of gnomAD (total allele count = 97, unique variants = 57; Chi Square test P-value for unique variants = 1.613e-05; Chi Square test P-value for total allele count = 0.001574). These two burden analyses show that, even though only two patients with *FZR1* variants were identified amongst 250 DEE patients within our research cohort, there are strong statistical data to support an association between rare *FZR1*-missense variants and DEE.

### DEE-associated *FZR1* variants lead to reduced^57^ protein abundance

FZR1 and its homologs in other species are highly conserved with *Drosophila* Fzr protein showing over 70.3% identity with human FZR1. The amino acids that are impacted by the missense variants that we identified in the patients are conserved in *Drosophila* Fzr (Figure 1A, S2A). To test the effect of the patient variants on protein stability and localization, we expressed FZR1 with an N-terminal eGFP tag in HEK293 cells. FZR1 expression was detected in the nucleus, with a weaker diffuse signal in the cytoplasm (Figure 1B). Variant FZR1 proteins showed a similar pattern of localization. However, western blot of total protein from cells transfected with wt or mutant eGFP-FZR1 showed a clear difference in FZR1 abundance, with both patient variants leading to a reduction in protein level of approximately 40% (p<0.01) (Figure 1C-D).

**Figure 1.**
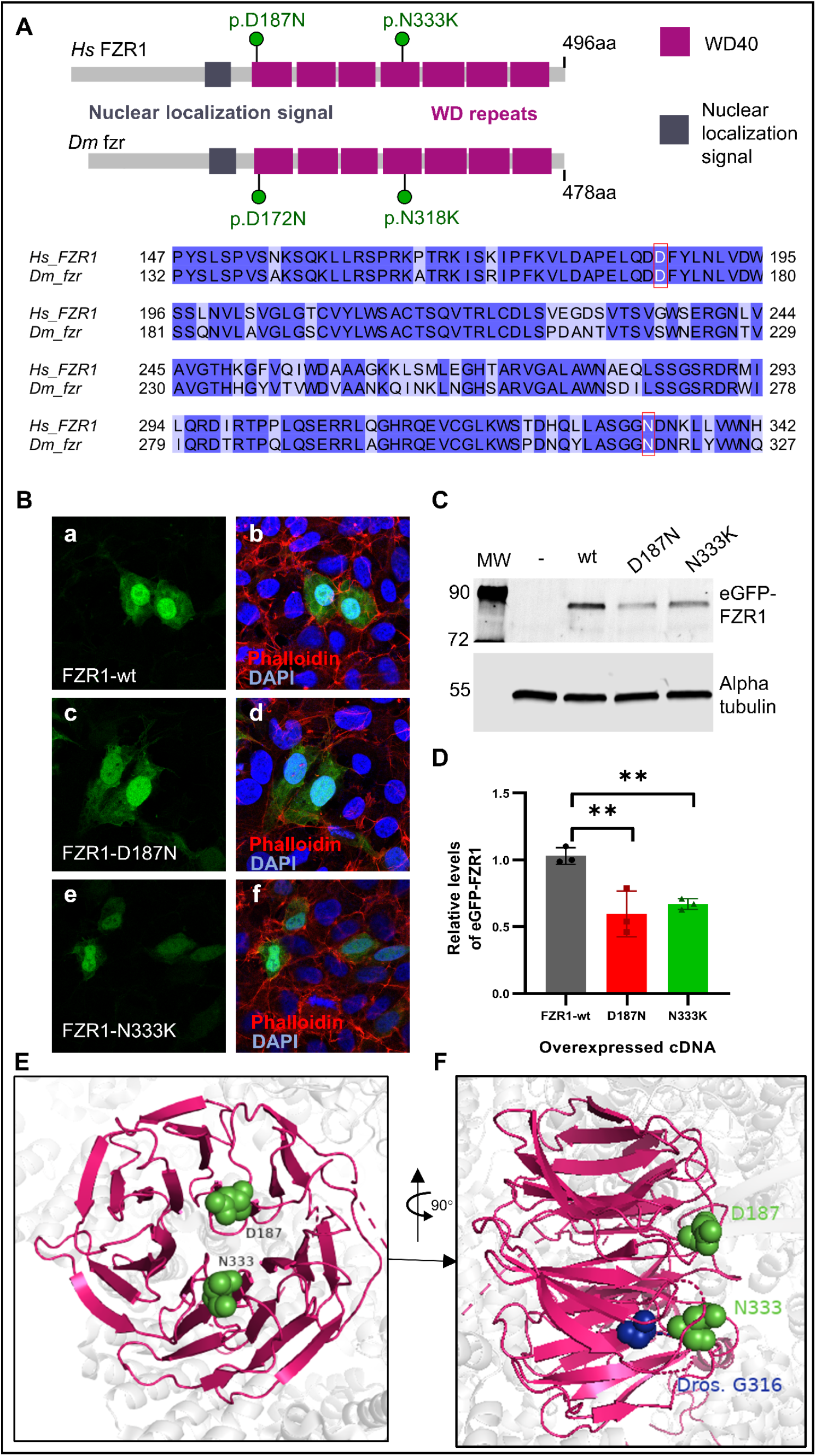
Conservation *FZR1* and in vitro analysis of DEE variants in mammalian cells. (A) Protein primary structure diagram of human FZR1 and *Drosophila* ortholog Fzr showing conserved domains and corresponding positions of variants observed in patients. The residues affected in the patients are conserved in *Drosophila melanogaster*. Amino acid sequence alignment of human FZR1 and *Drosophila* Fzr region encompassing the DEE variant residues which shown in red boxes. (B) Immunofluorescence staining for eGFP tagged human FZR1 cDNA in HEK293 cells (green), co-stained with DAPI (nucleus; blue) and actin-cytoskeleton (Phalloidin; magenta). (a-b) FZR1-wt, (c-d) FZR1:p.D187N, (e-f) FZR1:p.N333K localization to the nucleus and the cytoplasm. (C) Western blot showing relative expression of FZR1-wt and variants in HEK-293 cells. Alpha-tubulin is used as loading control. (D) Quantitation of normalized western blot signal analyzed using one-way ANOVA, followed by Dunnett test for comparison of the variants to the wt expression. ** indicates multiplicity adjusted P-value < 0.01. (E) 3D structural model FZR1 as part of the Cdh1-APC (PDB:4ui9)^9^ showing the relative positions of the variants affected in the DEE patients (green) and corresponding residue of the *Drosophila* mutation observed in the *fzr*^*B*^ allele (blue).

To further examine the effect of patient variants on human FZR1 function, we mapped the affected residues in a 3D protein structure of the APC^9^. Variants found in all three patients affect residues within the WD40 domain of the FZR1 protein (Figure 1E, S2B). WD40 domains, also known as beta transducing repeats, enable protein-protein interactions^58^, and *de novo* variants in another WD40 repeat containing protein encoding gene *WDR37* has also been recently associated with epilepsy^59^. The active site of a WD40 domain is often found in the central cleft of the propeller where the loops connect the successive beta sheets. As both variants are present in those loops (Figure 1D), they may interfere with the protein-protein interaction sites and potentially affect the substrate binding capacity of FZR1 in addition to reducing overall protein stability.

### Genetic characterization of *fzr* alleles in *Drosophila*

We decided to examine the functional effect of the DEE-associated variants that we identified using *Drosophila* as a model organism. For this study we used two EMS (ethylmethanesulfonate)-induced alleles of *fzr* (*fzr*^*A*^ and *fzr*^*B*^) that we previously isolated. These alleles produce rough eye phenotypes in adult mosaic animals and show defects in electroretinograms^22^, suggesting that *fzr* is involved in the development and function of the fly visual system. We determined the molecular lesions in these lines using Sanger sequencing of PCR fragments of genomic DNA. The molecular lesions in the *fzr*^*A*^ allele is a canonical splice-site mutation (fzr-RA:c.592+1G>A) and the *fzr*^*B*^ allele is a missense mutation (fzr-PA:p.G316N, Figure 2A). In the protein structure model, this fly mutation is found close to the p.N333K residue affected in Patient 2 and 3 (Figure 1F).

**Figure 2.**
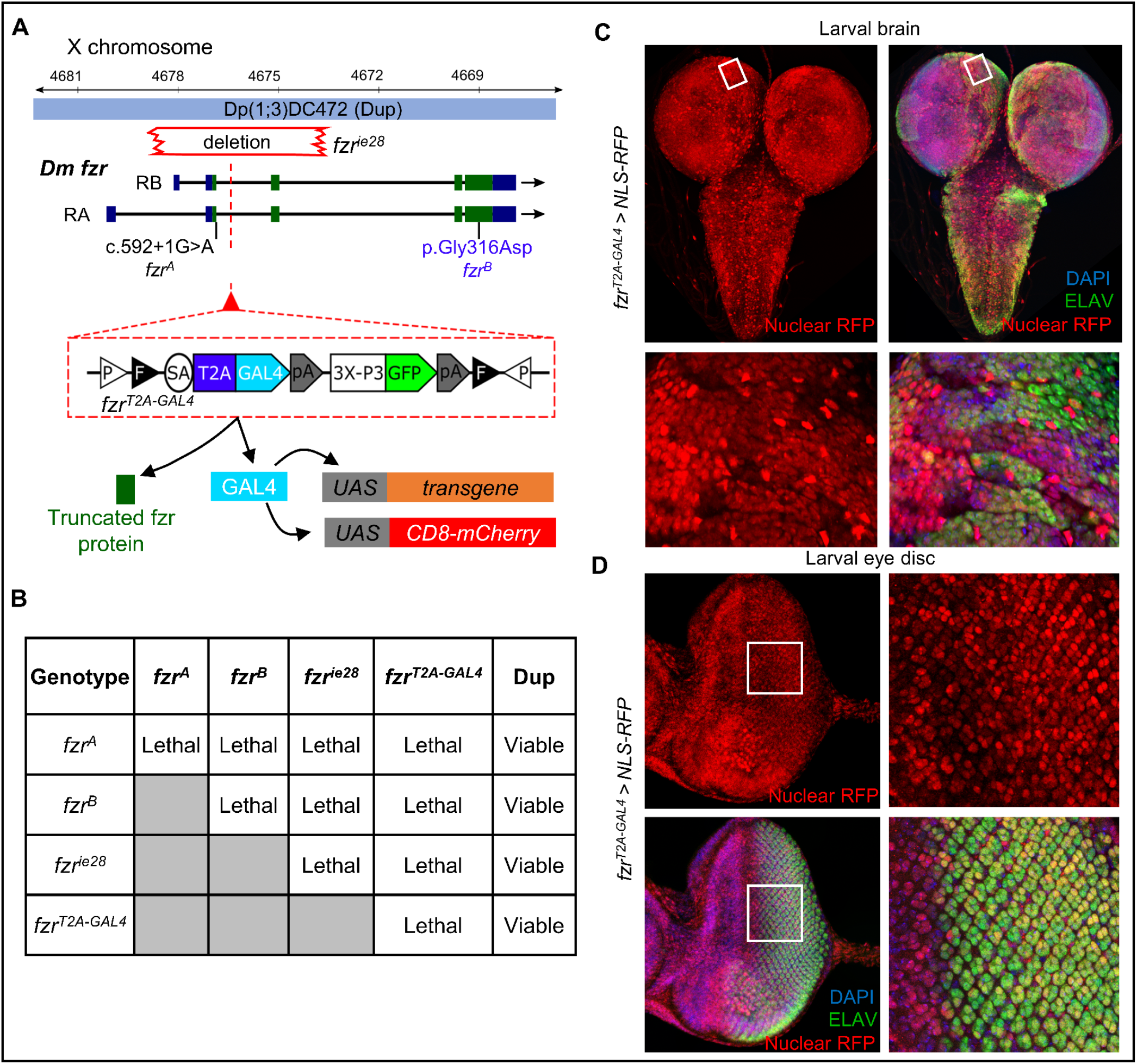
*Drosophila* fzr alleles and expression pattern. (A) *Drosophila X*-chromosome locus showing two transcriptional isoforms of *fzr*, two EMS-induced mutation alleles *fzr*^*A*^ (splice-site mutation) and *fzr*^*B*^ (missense mutation located in the same domain as one of the patient’s variants), *fzr*^*ie28*^ (deletion spanning the first two exons of *fzr*) and a third chromosome duplication allele carrying a complete copy of *fzr* from the *X-*chromosome. Also shown is the *fzr*^*T2A-GAL4*^ allele generated by insertion of a mutagenic T2A-GAL4 artificial exon within the second intron of *fzr*. This insertion leads to termination of *fzr* translation and expression of GAL4 protein from the endogenous *fzr* locus. (C) Table showing results of complementation tests between various alleles of *fzr* as well the *fzr* locus duplication shown in (B). (C-D) Expression pattern of fzr in *Drosophila* third instar visualized using the *fzr*^*T2A-GAL4*^ driving the expression of *nls::RFP* (red) co-stained with ELAV marking neuronal cells (green) and DAPI to detect nucleus (blue). (C) Larval brain and higher magnification images of the boxed region. (D) The eye-antennal disc and higher magnification images of the boxed region.

We created an additional fly mutant by using CRISPR/Cas9 and homology directed repair to introduce an artificial exon containing a splice acceptor-T2A-GAL4-polyA cassette between the first and second exons of *Drosophila fzr*. This allele is predicted to generate a strong LOF allele that also produces a GAL4 in the same spatial and temporal pattern reflecting the endogenous *fzr* expression pattern^25^. This CRIMIC insertion (*fzr*^*CR00643-TG4*.*2*^, henceforth called *fzr*^*T2A-GAL4*^) leads to precocious termination of transcription of all *fzr* isoforms because of the presence of a polyA termination signal as well as an interruption of translation of *fzr* mRNA due to the viral T2A peptide sequence. T2A also allows re-initiation of translation to express the GAL4 when and where endogenous *fzr* is expressed (Figure 2A). The *fzr*^*T2A-GAL4*^ allele permits expression of transgenes including *fzr* cDNA in the native pattern of *fzr* in the *fzr*-mutant background by crossing this line to a *UAS-fzr* transgenic fly (Figure 2A)^25^, simplifying the functional studies of the variants of interest.

We found that the *fzr*^*T2A-GAL4*^ allele is recessive lethal and fails to complement the lethality of a previously reported null allele of *fzr* (*fzr*^*ie28*^), suggesting that *fzr*^*T2A-GAL4*^ is a strong LOF allele (Figure 2B). Complementation test between the *fzr*^*T2A-GAL4*^ and the EMS-induced mutations (*fzr*^*A*^ and *fzr*^*B*^ ) reaffirmed that these mutations are allelic and are also LOF mutations (Figure 2B). Finally, we examined the rescue of the recessive lethality of *fzr*^*T2A-GAL4*^ and the EMS alleles using a duplication containing the *fzr* locus inserted on the third chromosome [*Dp(1;3)DC472, Dup*]. Each allele was individually rescued by this duplication and the rescued animals did not exhibit any morphological defects, confirming that the phenotypes observed in these alleles are due to LOF of *fzr* (Figure S3C). To examine the expression pattern of *fzr in vivo*, we crossed the *fzr*^*T2A-GAL4*^ allele to a *UAS-nls::RFP* transgenic line and explored the fluorescent pattern. We found that *fzr* is broadly expressed in larval brains and eye imaginal discs, consistent with their developmental roles in these tissues (Figure S3D-E).

### The role of *fzr* in neurodevelopment in embryos and larva

To further characterize the phenotypes of the *fzr* alleles we generated, we first examined the effect of the *Drosophila* EMS mutants in developing larval eye imaginal disc and adult retina. We tracked the stereotypical pattern of larval photoreceptor precursors in homozygous mutant cells induced through the expression of *eyeless* (*ey*) enhancer driven Flippase^57^ (*ey-FLP*) that is generated using the MARCM (Mosaic Analysis with a Repressible Cell Marker) technique^60^. In this experiment, Flippase mediates the recombination of two FRT (Flippase Recognition Target) sites located at the base of the X-chromosome (FRT19A). When this occurs during mitosis and one of the two sister chromatid carries the *frz* mutant allele, homozygous mutant and homozygous wild-type cells are generated from heterozygous cells (Figure 3A). Simultaneously, it reverses the repression of GAL4 by GAL80 in homozygous mutant cells, allowing the GAL4 to drive the expression of a GFP reporter and simultaneously an optional *UAS-fzr cDNA* transgene (wt or variant, Figure 3B). Consequently, mutant cells are marked by the GFP reporter and if included, transgenes are expressed within these mutant cells to rescue the phenotype caused by *fzr* LOF.

**Figure 3.**
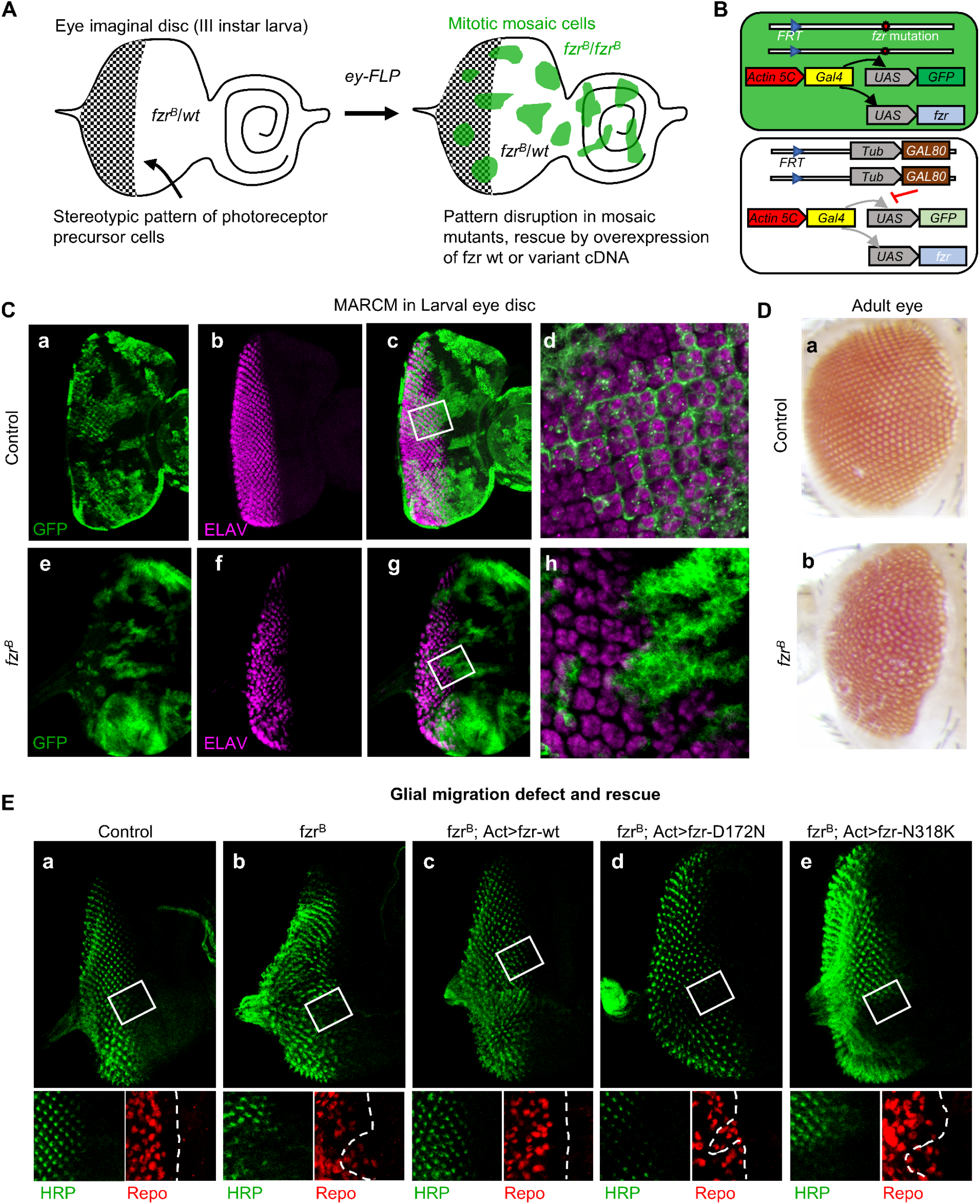
Pattern formation and glial migration defects in the photoreceptor precursors of *fzr* mutants are not rescued by DEE variants. (A) Diagram showing the generation of random mosaic patches of homozygous *fzr* mutant cells in the developing eye disc that are marked by GFP expression using MARCM. (B) Schematic diagram showing gene expression differences between homozygous *fzr* mutant (GFP+) cells and the surrounding heterozygous *fzr* (GFP-) cells. In mutant cells, *Actin5c-GAL4* drives the expression of GFP and *fzr* cDNA (wild-type or patient variant), while in heterozygous cells, GAL4 activity is suppressed by GAL80. (C a-d) Control larval eye discs showing expression pattern of ELAV (red) in mosaics marked by GFP expression generated using MARCM. (C e-h) larval eye disc showing mosaic homozygous *fzr*^*B*^ tissue (green) showing dramatic change in ELAV (magenta) expression within and outside the clones, showing cell autonomous and non-autonomous effect of *fzr* loss *in vivo*. (D-a) Control adult eye showing stereotypical pattern of compound eye. (D-b) Aberrant retina pattern observed in *fzr*^*B*^ mosaic adults. (E) MARCM mosaics generated as above stained with HRP (photoreceptor membrane). Magnified regions showing HRP boundary in dotted lines and glial cells stained with ani-Repo antibody. (E-a) Control larval eye imaginal disc showing limitation of glial cells to the boundary set by HRP positive cells (boundary traced by white dotted lines). (E-b) *fzr*^*B*^ mosaics showing not only disturbances in HRP pattern but also migration of glial cells past the boundaries (arrowhead) of HRP positive cells. (E-c) Eye imaginal discs showing rescue of HRP pattern and glial cell migration when *fzr* wild-type cDNA is overexpressed in mutant mosaics. (E-d) Eye imaginal discs showing glial migration in regions lacking HRP signal when *fzr* cDNA with variant p.D172N is overexpressed in *fzr* mutant cells. (E-e) Eye imaginal discs showing similar defects in glial cell migration when *fzr* cDNA with variant p.N318K is expressed in *fzr* mutant cells.

In the larval eye disc, the developing photoreceptors were marked by staining for ELAV (Embryonic lethal abnormal vision), a pan-neuronal nuclear marker^61^. In the homozygous mutant cells identified using GFP reporter, we found that the larval photoreceptor patterns marked by ELAV were severely affected (Figure 3C). This phenotype in the *fzr*^*B*^ allele is consistent with previous reports of *fzr* function in the eye^17^. In adults the pattern of the compound eye was also significantly affected in animals in which mutant mosaic clones were generated (Figure 3D).

To further assess the function of photoreceptors, we performed ERG recordings on 3-4-day old flies. Adult flies with *fzr* mosaic eyes showed a significant reduction in the depolarization amplitude of the ERG response to light (Figure S2A-B), indicating a defect in phototransduction. These flies also showed severe loss of the ON/OFF transients (Figure S2A-B), suggesting a defect in synaptic transmission.

Next, we examined the embryonic neurogenesis phenotype of the *fzr*^*T2A-GAL4*^ mutants by assessing the morphology of the nervous system through immunofluorescence staining and confocal microscopy using an antibody that recognizes neuronal/photoreceptor membranes (anti-HRP antibody). Hemizygous *fzr*^*T2A-GAL4*^ mutant males showed a clear defect in neuronal patterning in the central nervous system of fly embryos, displaying defective neuromere patterns in the ventral nerve chord (Figure S2C). This is consistent with the role of *fzr* in neuronal development and phenotypes reported for previously identified *fzr* alleles^19^.

### DEE-associated variants fail to support *Drosophila* neurodevelopment in rescue experiments *in vivo*

*fzr*^*B*^ mosaic flies showed severe morphological defects in larval eye imaginal discs as evidenced from aberrant anti-HRP staining pattern in the mutant mosaic clones (Figure 3E). We examined the ability of the *fzr* cDNA with wt sequence or cDNAs with variants corresponding to those of the human patients (Figure 1A) to rescue this phenotype MARCM)^62^. We observed that overexpression of the wt fly fzr cDNA in the mutant mosaic clones rescued the HRP pattern defects in the eye discs (Figure 3E-c), but the variant cDNAs failed to display a similar rescue (Figure 3E-d,e).

*fzr* mutants in *Drosophila* have also been reported to exhibit defects in glial cell migration^17^. Since glia are known to play important roles in epilepsy^63^, we examined the impact of the two patient variants on this phenotype, again using the MARCM system in the eye imaginal disc. In a control animal (mosaic animals without *fzr* mutant tissue, Figure 3E-a), glial cells only migrate to areas where photoreceptors have initiated their differentiation program indicated by HRP-positive cells (arrowhead, Figure 3E-a). However, in mosaic cells that are defective for *fzr*, we observed that glial cells migrate beyond the differentiated zone and prematurely enter areas where immature photoreceptors are present (HRP negative region, Figure 3E-b). We observed that wt *fzr* cDNA was able to suppress this phenotype (Figure 3E-c) but the *fzr* cDNAs carrying the patient’s variants were not able to rescue the abnormal migration of glial cells (Figure 3E-d,e). This suggests that neuron-glia communication mediated by *fzr* are also affected by the patient variants.

Since we noticed that overexpression of *fzr wild-type* cDNA in the eye clones occasionally led to aberrant development patterns, we examined if overexpression of *fzr* cDNA in wild-type background can disrupt the normal photoreceptor development pattern. Indeed, when wt *fzr* cDNA was overexpressed in the developing eye using *ey-GAL4*, we observed a severe reduction in the size of adult retina and loss of photoreceptor pattern (Figure 4B), similar to what has been reported earlier ^21^. In this assay, we observed that *fzr* cDNA with patient variants also showed caused aberrant photoreceptor patterns (Figure 4C-D). However, the reduction in the size of the adult retina due to the *fzr* variant overexpression was not as severe as the *fzr* wt cDNA and the differences between the overexpression of wt and variants were statistically significant (Figure 4E). These results further support that *FZR1* variants associated with DEE are loss of function alleles.

**Figure 4.**
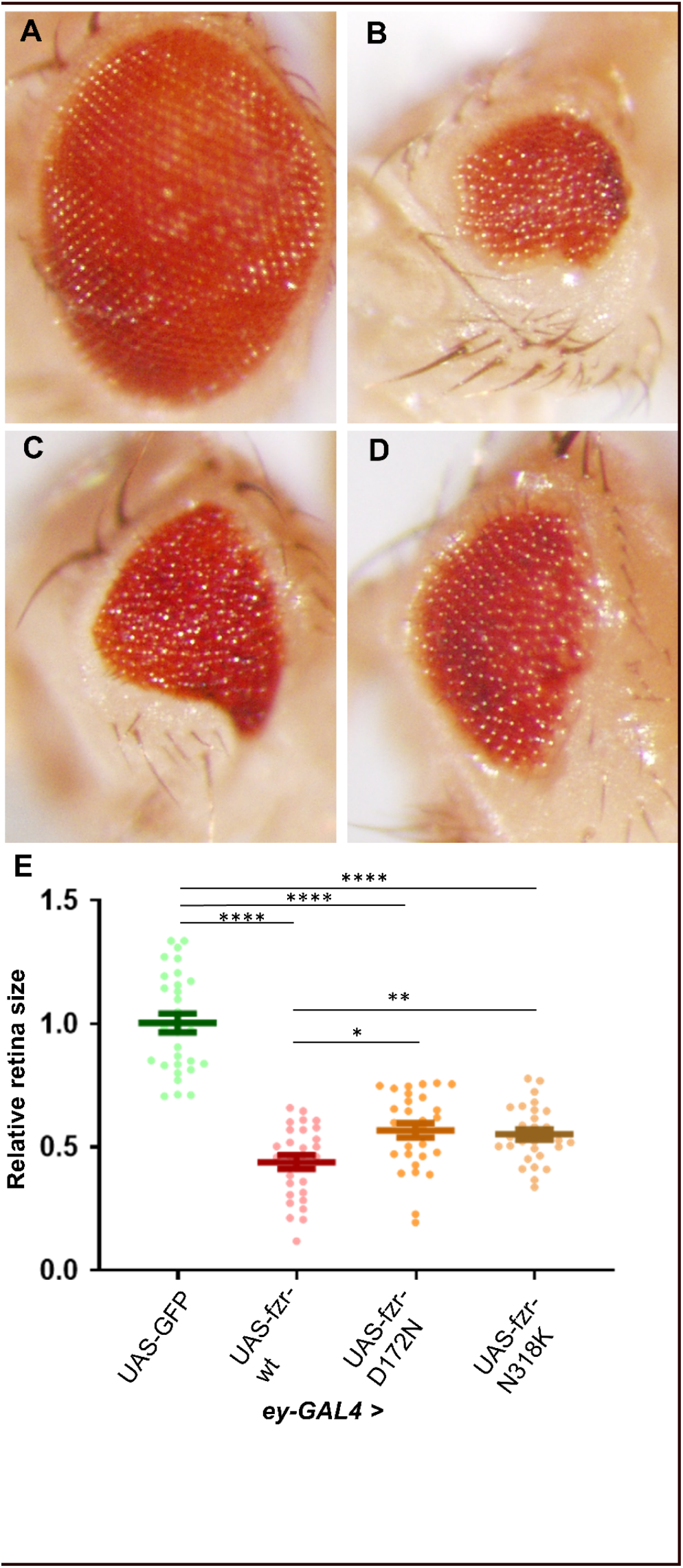
Overexpression of *fzr* leads to aberrant retinas. (A) Control *ey3*.*5-GAL4* eyes showing stereotypical photoreceptor pattern and retina size. (B) eyes of animals with wild-type *fzr* overexpressed using *ey3*.*5-GAL4* showing severe eye size reduction and disrupted ommatidia pattern. (C) Adult eyes of *ey3*.*5-GAL4* mediated overexpression of Fzr:p.D172N leading loss of photoreceptor pattern and reduction in retina size. (D) Adult eyes *ey3*.*5-GAL4* mediated overexpression of Fzr:p.N318K leading to disruption in ommatidia pattern and reduction in eye size. (E) Quantitation of the eye size shows significant reduction of retina area due to the overexpression of wild-type or variant *fzr* cDNAs. Reduction in the eye size due to variant overexpression is not as severe as defects caused by wild-type Fzr and are statistically significant. Statistical difference evaluated using one-way ANOVA between all the samples followed by pair-wise analysis using Tukey’s multiple comparison. (multiplicity corrected P-value indicated as **** < 0.0001, ** < 0.001, * < 0.05).

We next examined if wild-type *UAS-fzr cDNA* can rescue the lethality observed in *fzr*^*T2A-GAL4*^ flies. We used GAL4 expressed from the T2A-GAL4 allele to drive the expression of the *UAS-fzr*. Expression of wild-type *fzr* cDNA was able to allow a significant fraction (∼20-23%) of *fzr*^*-T2A- GAL4*^ hemizygous males to live to third instar larval stages whereas they otherwise die as embryo or first instar larvae (Figure 5A). In contrast, the overexpression of *fzr* cDNA carrying either of the two patient variants fails to rescue this early lethality (Figure 5A). Finally, we examined whether the patient *fzr* cDNA can rescue the central nervous system development defects in *the fzr*^*T2A-GAL4*^ mutant embryos. The wild-type *fzr* cDNA was able to restore this defect (Figure 5D), but the two variant cDNAs did not rescue the lesions (Figure 5E-F). Together, the MARCM-based rescue experiments in the developing eye, overexpression analysis in the developing eye, and rescue studies of embryonic neuronal developmental defects all suggest the variants found in three DEE patient act as strong hypomorphic alleles, impacting neuronal development in multiple contexts.

**Figure 5.**
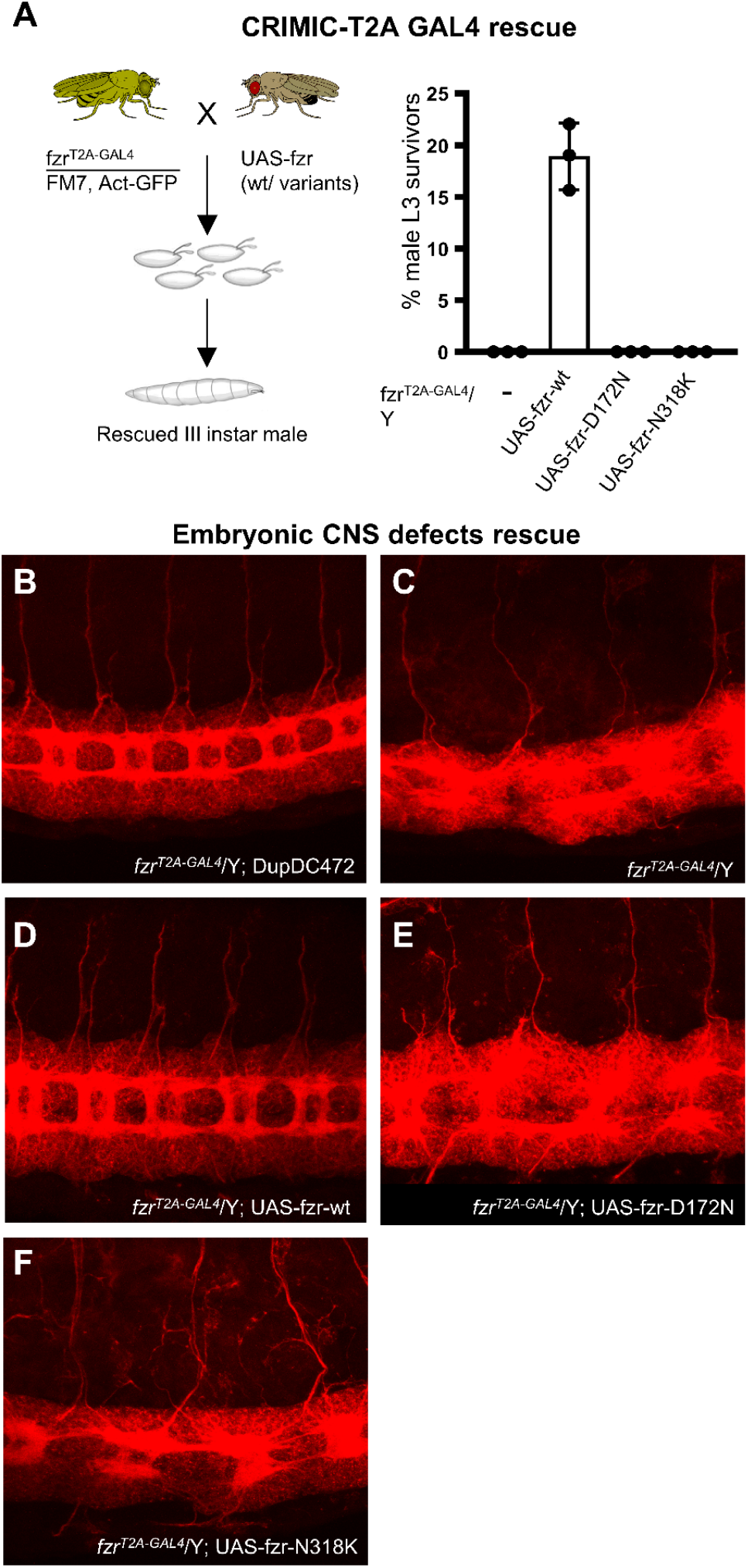
Embryonic central nervous system defects in *fzr*^*T2A-GAL4*^ mutants are not rescued by Fzr carrying DEE patient variants. (A) Diagram showing the crossing scheme used to assess rescue of lethality observed in *fzr*^*T2A-GAL4*^ using overexpression of wt or variant *Drosophila* Fzr and bar plots showing percentage of later third instar (L3) male larvae recovered by the overexpression of wild-type and variant cDNAs in a *fzr*^*T2A-GAL4*^ mutant background. (B-F) *Drosophila* embryos at stage 15-16 showing neuronal membranes stained with HRP. (A) *fzr*^*T2A-GAL4*^ hemizygous embryo (*fzr*^*T2A-GAL4*^ */Y*) rescued with duplication of *fzr* genomic region on third chromosome serving as control. (B) *fzr* ^*T2A-GAL4*^ /Y embrryos showing severe defects in the patten of neuron development. (C) Wild-type Fzr expression using the *fzr*^*T2A-GAL4*^ rescues neuronal pattern. (D-E) *fzr*^*T2A-GAL4*^ shows neurodevelopmental defects that are not rescued by the expression of Fzr p.D172N or p.N318K.

## Discussion

In this study, we describe three individuals with DEE with childhood onset generalized epilepsy carrying a *de novo* missense variant in *FZR1*. One missense variant affects the same amino-acid residue as identified in a single previously published patient with DEE^24^. We further provide statistical and functional support for pathogenicity of these variants.

All four individuals carrying pathogenic *de novo FZR1* variants described so far suffer from neurodevelopmental delay and epilepsy. While the previously reported individual had neonatal onset treatment-resistant multifocal seizures, severe ID and prenatal microcephaly^24^, our study shows that the phenotypical spectrum also includes childhood onset generalized seizure syndromes associated with moderate to severe ID, mild ataxia and normal head circumference. This variability could be due to strength of alleles or genetic backgrounds, which will require further investigation. Of note, two patients in our study were diagnosed with MAE. A normal development prior to seizure onset is historically considered a diagnostic criterium for the diagnosis of MAE, but the phenotypic boundaries of this syndrome remain debated^64^. In a recent study on the genetic etiology of MAE, more than 20% of patients did have developmental delay prior to seizure onset^8^. This feature is indeed inherent to the concept of DEE, which acknowledges that the neurodevelopmental impairment of these patients is not solely related to frequent epileptic activity but is also a direct result of the underlying gene dysfunction. Of note, the <5 years-old patient 3 in our study did not have (myoclonic) atonic seizures at time of inclusion in this study. As early disease history is very similar to patient 2, carrying a variant leading to the same amino-acid substitution, further clinical evolution will tell whether other seizure types will still occur. Both patients also had similar signs of delayed myelination on early brain MRI.

There is a significant enrichment of both *de novo* and of (predicted) deleterious variants in our patient cohort of DEE patients compared to a control population such as gnomAD. Interestingly, all four DEE associated variants described so far affect one of two different residues of FZR1. *FZR1* variants found in DEE patients affect residues that are likely to be important for the substrate recognition of Cdh1-APC^9^. Variants in this region may lead to altered substrate recognition and therefore lead to a diminished function of Cdh1-APC. Furthermore, we have shown that our patient variants lead to lower protein levels, which could indicate a reduction in the stability of mutant FZR1.

Using our *Drosophila* functional assays, we show that both p.D187N and p.N333K variants lead to functional deficits in Fzr (Cdh1) protein, especially in the developing nervous system. The DEE-associated variants fail to rescue photoreceptor pattern and glial cell migration that are phenotypes observed in previous *fzr* alleles in *Drosophila* and the patient variants behave as partial loss of function alleles. This is also observed in an overexpression assay in the developing eye, as we find that the variants display retention of some function compared to the reference allele. The difference in the functionality between the variants and the wt Fzr is dramatic in the embryonic CNS development as observed using the *fzr*^*T2A-GAL4*^ allele, which might provide a more sensitive readout of functional differences. As a regulatory subunit of the E3 ubiquitin ligase complex APC, FZR1 (Cdh1) is involved in the turnover of many substrates^65^. One substrate of Cdh1-APC is FMRP (Fragile X mental Retardation Protein) encoded by the *FMR1* (MIM:309550) gene^66^. The interaction of FMRP with Cdh1-APC was shown to regulate metabotropic glutamate receptor (mGluR)-dependent synaptic plasticity^66^ and the formation of stress granules and protein synthesis-dependent synaptic plasticity^67^. Triplet repeat expansion in *FMR1* causes Fragile X syndrome, a neurodevelopmental disorder often accompanied by seizures^68^. HECW2 (HECT, C2 And WW Domain Containing E3 Ubiquitin Protein Ligase 2, also known as NEDL2 MIM:617245) is another substrate of Cdh1-APC^69^ with a link to neurodevelopmental disorders, as *de novo* missense variants lead to ID, seizures and absent language^70^. Cdh1-APC-mediated degradation of HECW2 during mitotic exit is important for the regulation of metaphase to anaphase transition^69^. Abnormality of these or other substrates of Cdh1-APC may underlie the DEE and other phenotypes seen in our patients, which will require further molecular studies.

In summary, our work provides genetic, statistical and functional support for the role of *FZR1* in DEE, and we expand the phenotypic spectrum of *FZR1*-related encephalopathy to include individuals with DEE with childhood onset generalized epilepsy and normal head circumference. The presence of early neurodevelopmental delay, even prior to seizure onset, is in line with the deleterious impact of patient variants on early neurodevelopment in a *Drosophila* model. The three molecularly defined *fzr* alleles (*fzr*^*A*^, *fzr*^*B*^, *fzr*^*T2A-GAL4*^) are of great value to the study of the role of *fzr* in *Drosophila*, and due to the extensive conservation of residues, the study of *FZR1* in human diseases. The fly mutant, overexpression lines, and assays that we have developed in this study will be an important asset for future studies on the role of *FZR1* in neurodevelopment and the impact of human disease variants on its interaction with substrates such as FMRP and HECW2 in the development of DEE.

## Data Availability

All relevant data and supporting information are contained within the manuscript and supporting data files.

https://gnomad.broadinstitute.org/

https://denovolyzer.org/

https://www.ensembl.org/info/docs/tools/vep/index.html

http://www.omim.org/.

https://www.rcsb.org/

## Supplemental Information

Supplemental data include three figures and one table.

## Acknowledgements

N.S. is supported by the UA-Bijzonder Onderzoeksfonds (BOF)-DOCPRO4 (FFB180186).

S.Y. is supported by the following grants from the National Institutes of Health (U54NS093793, R01DC014932, R24OD022005) and through funds provided by the Nancy Chang Ph.D., Award for Research Excellence, Baylor College of Medicine, and the Jan and Dan Duncan Neurological Research Institute at Texas Children’s Hospital. S.W. is supported by the Fonds Wetenschappelijk onderzoek (FWO 1861419N). H.M. is supported by NIH (R01 NS069605). We would like to thank Dr. Michael Wangler for guidance on the bioinformatic analysis of the variants and comments and discussion on the manuscript.

## Web resources

gnomAD https://gnomad.broadinstitute.org/

denovolyzeR https://denovolyzer.org/

Variant Effect Predictor https://www.ensembl.org/info/docs/tools/vep/index.html

OMIM http://www.omim.org/.

PDB https://www.rcsb.org/

## Data and Code Availability

All WES data generated in the EuroEPINOMICS-RES project was deposited in the European Genome-phenome Archive, accession numbers EGAS00001000190, EGAS00001000386, and EGAS00001000048.

