## Supplemental Information for "*De novo FZR1* loss-of-function variants cause developmental and epileptic encephalopathies including Myoclonic Atonic Epilepsy"

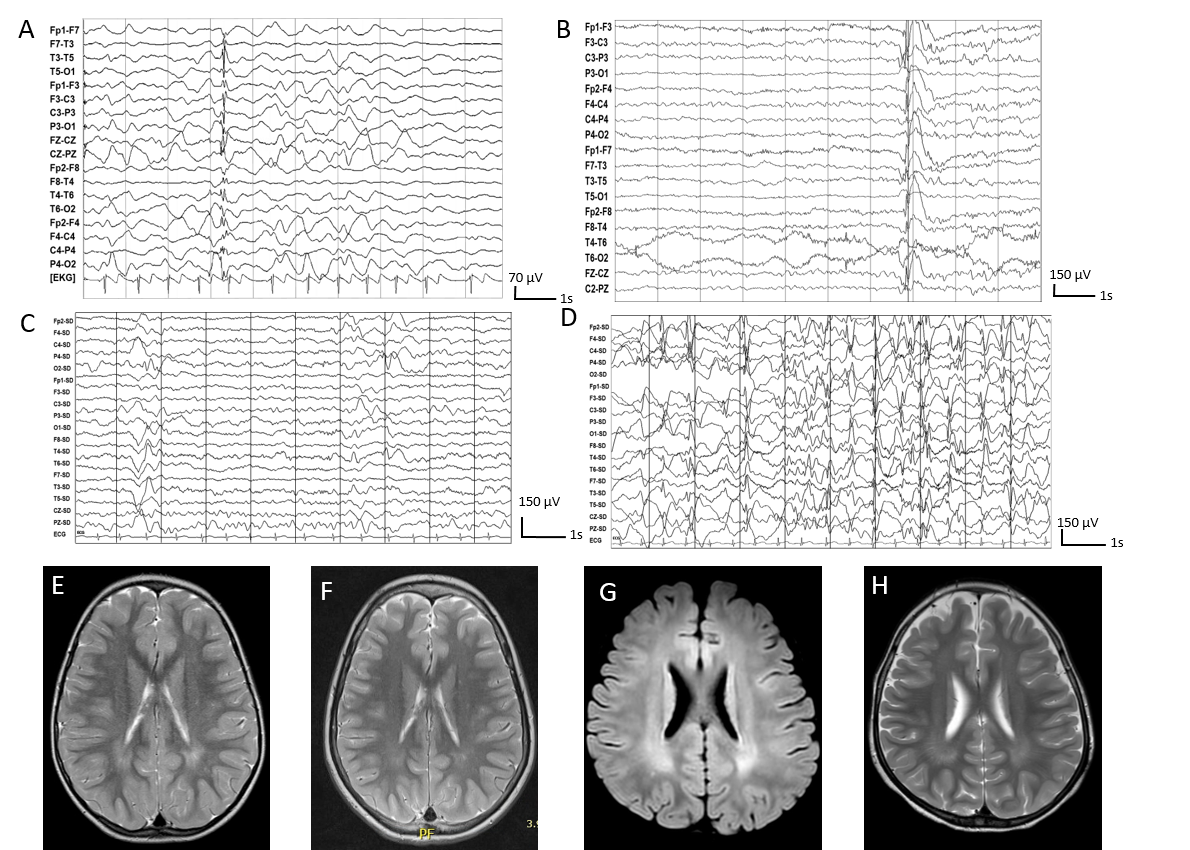

### **Supplemental Information**

#### Figure S1. Imaging of DEE patients with FZR1 variants. (A) EEG recording during sleep of patient 1, showing an isolated generalized polyspikewave. (B) Awake EEG of patient 2 showing an isolated generalized spike wave. (C-D) Awake EEG of patient 3 showing a slow background and bursts of generalized sharp waves and irregular slow spike-wave activity. (E) Axial T2 weighted MRI brain image of patient 2 showing bilateral parieto-occipital white matter hyperintensities. (F) Axial T2 weighted MRI brain image of patient 2 again showing an almost complete disappearance of white matter hyperintensities. (G-H) Axial FLAIR and T2 weighted MRI of patient 3, showing mild frontotemporal accentuated cerebral atrophy and bilateral white matter hyperintensities. Precise age of the patient at the time of capture of these images can be obtained upon a reasonable request from the corresponding author.

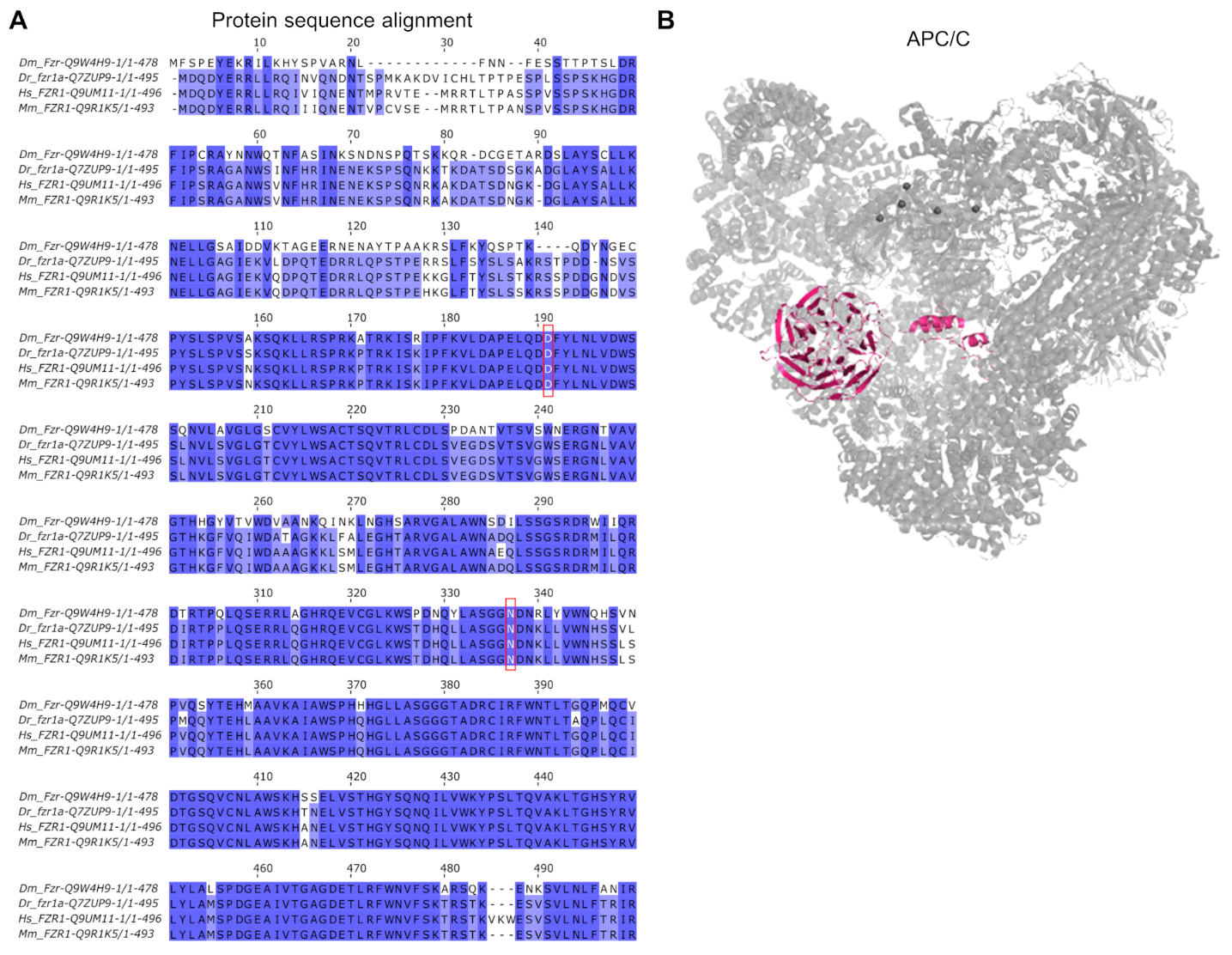

#### **Figure S2. Conservation of FZR1 protein and the expression of *Drosophila* *fzr* gene.**

(A) ClustalOmega based amino acid conservation of human (Hs), mice (Mm), zebrafish (Dr), and *Drosophila* (Dm), showing conserved residues highlighted in blue. Residues affected in DEE patients are highlighted in red boxes. (B) 3D structural model (PDB:4ui9) of Cdh1-APC showing the relative position of FZR1 in the complex.

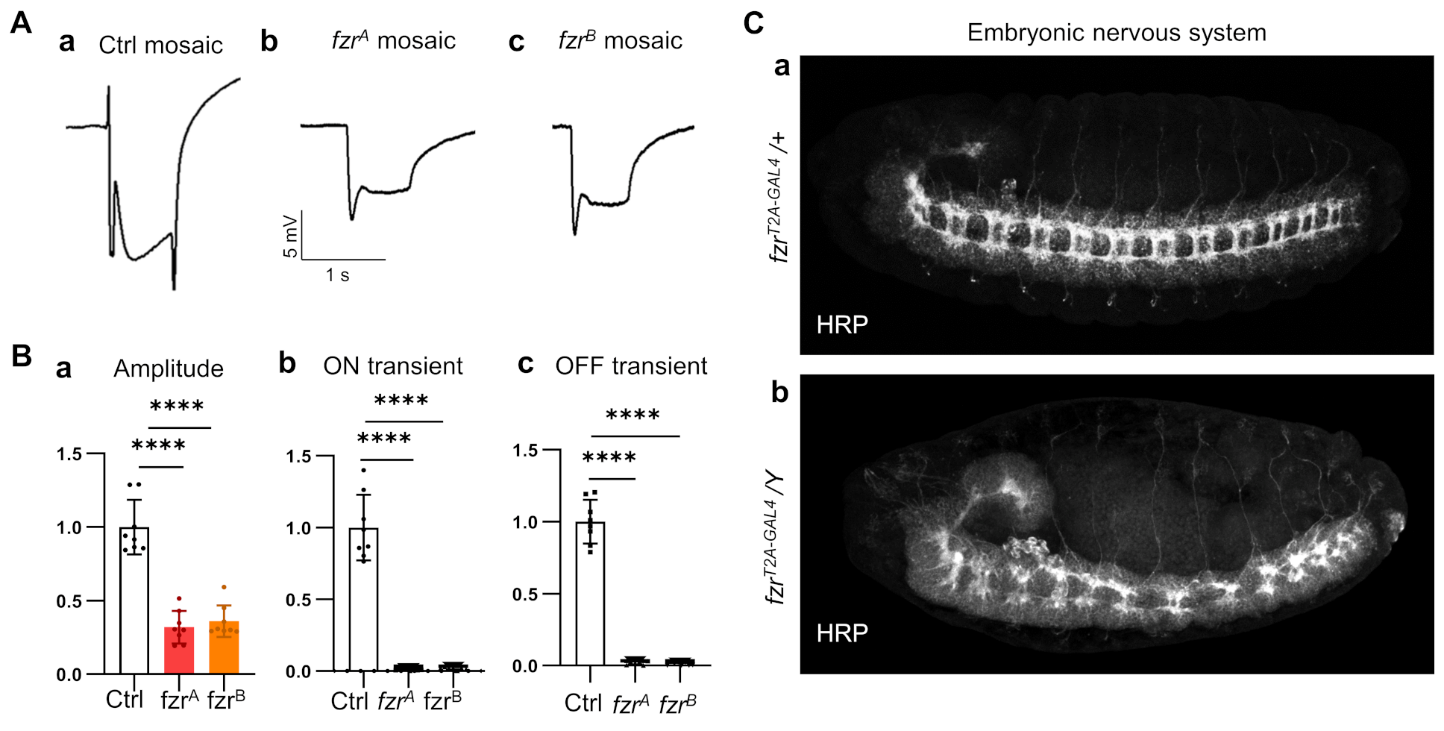
**Figure S3. Neuronal phenotypes observed in the *Drosophila* *fzr* alelles:** (A) Electroretinograms of adult *Drosophila* eyes from mosaic eyes of (a) control [isogenic *y w FRT19A* chromosome clones], (b) *fzr^A^* and (c) *fzr^B^* animals. (B) Quantification of the depolarization amplitude (a), and ON (b) or OFF (c) transients from the electroretinogram (n=8/genotype). (C-a) HRP staining of a fly embryo heterozygous for *fzr^CRIMIC-T2A-GAL4^* showing stereotypical pattern of *Drosophila* central nervous system in stage 15-16 embryo. (C-b) HRP stained *Drosophila* *fzr^T2A-GAL4^* hemizygous male embryos showing severe defects in the pattern of central nervous system.

#### Supplemental Table 1.

MAE-related genes and candidate genes included on the MAE-panel for patient 2 (in alphabetical order).

| **HGNC gene name** | **MIM** |
| --- | --- |
| *ABHD15* | NA |
| *ANLN* | 616027 |
| *ANO4* | 610111 |
| *AP2M1* | 601024 |
| *ARSA* | 607574 |
| *ATXN2L* | 607931 |
| *BAZ2A* | 605682 |
| *C12orf50* | NA |
| *CAP2* | 618385 |
| *CCDC141* | 616031 |
| *CDK16* | 311550 |
| *CHD2* | 602119 |
| *CSNK2A1* | 115440 |
| *ELL2* | 601874 |
| *EPC2* | 611000 |
| *FZR1* | 603619 |
| *GABRA1* | 137160 |
| *GDI2* | 300104 |
| **HGNC gene name** | **MIM** |
| *GINM1* | NA |
| *GLI3* | 165240 |
| *GPHN* | 603930 |
| *GXYLT2* | 613322 |
| *HCN1* | 602780 |
| *KCNA2* | 176262 |
| *KRT78* | 611159 |
| *LCOR* | 607698 |
| *LCP1* | 153430 |
| *LIPK* | 613922 |
| *LTBP4* | 604710 |
| *NBEA* | 604889 |
| *NEXMIF* | 300524 |
| *PCDH19* | 300460 |
| *PPP4R2* | 613822 |
| *RANGRF* | 607954 |
| *RPTOR* | 607130 |
| *RTF1* | 611633 |
| **HGNC gene name** | **MIM** |
| *SCN1A* | 182389 |
| *SCN1B* | 600235 |
| *SCN2A* | 182390 |
| *SCN8A* | 600702 |
| *SLC25A12* | 603667 |
| *SLC35A4* | NA |
| *SMARCA2* | 600014 |
| *SNTB2* | 600027 |
| *SPIN1* | 609936 |
| *STX1B* | 601485 |
| *SYNGAP1* | 603384 |
| *TRPM1* | 603576 |
| *URB2* | NA |
| *ZNF483* | NA |
| *ZNF547* | NA |
| *SLC6A1* | 137165 |
